# Leveraging Large Language Models to Develop an Interpretable Prediction Model for Postpartum Hemorrhage Prior to the Onset of Labor

**DOI:** 10.1101/2025.03.23.25324452

**Authors:** Elizabeth G Woo, Israel Zighelboim, Tyler Gifford, Joseph G Bell, Hannah Milthorpe, Emily Alsentzer, Ryan E Longman, Jorge E Tolosa, Brett K Beaulieu-Jones

**Affiliations:** Center for Computational Medicine and Clinical AI, Department of Medicine, University of Chicago, Chicago, IL; Department of Obstetrics and Gynecology, St. Luke’s University Health Network. Bethlehem, PA; St. Luke’s Cancer Center, Division of Gynecologic Oncology, St. Luke’s University Health Network. Bethlehem, PA; Department of Biomedical Data Science, Stanford University, Stanford, CA; Department of Obstetrics and Gynecology, Section of Ultrasound, Genetics, and the Fetal Neonatal Care Center, University of Chicago, Chicago IL; Oregon Health and Science University, Maternal Fetal Medicine, Portland, Oregon

## Abstract

**Objective:** To evaluate whether large language models (LLMs) applied to prenatal clinical notes can predict postpartum hemorrhage (PPH) prior to the onset of labor and to compare model performance across outcome definitions, including a novel intervention-based definition.

**Methods:** We conducted a retrospective cohort study of 19,992 deliveries within a large regional health network. Two outcome definitions for PPH were used: estimated or quantitative blood loss (EBL/QBL) extracted from clinical notes, and a clinical intervention-based definition (cPPH) incorporating transfusion, uterotonics, Bakri balloon, or hysterectomy. We evaluated three approaches for PPH prediction: (1) supervised machine learning using structured electronic medical record data; (2) direct prediction using a fine-tuned LLM applied to clinical notes; and (3) interpretable models using LLM-extracted features combined with structured data. Model performance was evaluated using area under the receiver operating characteristic curve (AUROC) on a temporally held-out test set.

**Results:** The LLM-based direct prediction model achieved the highest performance for both PPH definitions (AUROC 0.79–0.80), followed by interpretable models combining LLM-extracted features with structured data (AUROC 0.76–0.78). Models using only structured data performed worse (AUROC 0.65–0.71). The LLM-extracted features approach identified 47 significant predictors, including established risk factors such as multiple gestation and previous cesarean delivery. Demographic differences were observed between PPH definitions: mothers who met only the cPPH definition had lower gestational age and higher rates of cesarean delivery compared to those meeting only the EBL/QBL definition.

**Conclusion:** These findings highlight the potential of LLM-based approaches for enhancing PPH risk stratification, with the feature extraction method offering a promising balance between predictive performance and clinical utility. Integrating these methods into clinical workflows could improve early detection and guide targeted preventive interventions.

## INTRODUCTION

Postpartum hemorrhage (PPH) is a leading cause of maternal morbidity and mortality, accounting for over 25% of maternal deaths worldwide.^1^ Prevention of serious complications depends on early recognition, resource availability, and prompt intervention. Management and prevention measures could be improved through accurate risk stratification and prediction models that enabling earlier identification of those at high risk for PPH. Several PPH risk stratification tools exist in the United States, including the California Maternal Quality Care Collaborative (CMQCC)^2^, Association of Women’s Health, Obstetric and Neonatal Nurses (AWHONN)^3^, and New York Safety Bundle for Obstetric Hemorrhage (NYSBOH)^4^. These tools rely on known, predefined risk factors including low platelet count, multiple gestation, prior cesarean delivery, and prior history of PPH. While these tools provide a structured framework for identifying high-risk individuals, they have demonstrated limited clinical utility^5,6^, motivating the development of novel models to accurately predict PPH risk.^7^ Recent efforts, such as the antepartum screening tool implemented in Epic at North Memorial Health, have demonstrated real-world success with a reduction in PPH rates.^8^

Early PPH prediction models used statistical models including logistic regression with variable performance.^9^ For example, a 2014 study using data from the Hypertension and Preeclampsia Intervention Trial At Term (HYPITAT) trial in the Netherlands failed to perform accurate antepartum prediction using logistic regression and achieved only modest performance with the addition of variables from the delivery encounter.^10^ More recent approaches have incorporated machine learning methods such as those used by Venkatesh et al. in 2020. This study found that while both machine learning and statistical models could accurately predict PPH, models including extreme gradient boosting and random forests achieved better performance than logistic regression models, though at the cost of increased complexity.^11^ Many existing models rely heavily on clinical and physiologic data collected during the delivery admission, limiting their capability to preemptively identify individuals at risk of PPH.^12^ Recent efforts have been made to develop models using data available upon labor admission or prior to delivery in order to improve prospective evaluation.^7,11,13,14^

The majority of predictive models for PPH rely solely on structured EMR data, which consist of variables such as laboratory results, medication orders, vital signs, and diagnosis codes. This could miss additional context that may be better captured in unstructured data such as clinical notes. Clinical information extraction from unstructured notes can be automated using large language models (LLMs), which are artificial intelligence (AI) systems trained on vast amounts of data that can be used to process and generate text. LLMs have contributed to a new era of natural language processing and have demonstrated the ability to process unstructured data within clinical notes. Notably, they are capable of extracting clinically relevant information without the need for additional labeled data, both in general contexts^15–17^ and in the context of postpartum hemorrhage.^18^ They have also been used to quantify the level of predictive signal in clinical notes for outcomes such as seizure recurrence, substantially outperforming prediction models which use structured data from electronic medical records (EMRs).^19^ LLMs have been used for direct prediction tasks in medical contexts such as readmission, mortality, length of stay, and comorbidity prediction.^20^ A challenge with LLM-based clinical risk prediction, however, is a lack of interpretability as it can be difficult to trace how predictions are made. A more interpretable method could use LLMs for intermediate feature extraction, as described in McInerney et al.^21^ These intermediate features can be manually validated before being used as input to models for downstream predictive tasks, resulting in more interpretable predictions.

An effective prediction model for PPH should not only be interpretable but also have a well-defined outcome of PPH. Multiple definitions are in use worldwide, most relying on estimates or measurement of blood loss, with or without consideration of other clinical parameters.^22^ The traditional definition of PPH was based on blood loss thresholds and mode of delivery.^23^ For example, the California Quality Care Collaborative suggested implementation of a PPH protocol for blood loss ≥ 500 mL for vaginal births and ≥ 1000 mL for cesarean births.^24^ Definitions of PPH based on estimated blood loss (EBL) can be suboptimal as estimated blood loss assessment is often inaccurate and blood loss at the time of delivery is routinely underestimated.^22,23,25^ While methods such as volumetry and gravimetry for quantitative measurement of blood loss alleviate some challenges associated with the more common method of visual estimation, attempts to implement these quantitative methods in Labor and Delivery units have been inconsistent.^26–28^ Additional definitions have sought to incorporate clinical signs and symptoms. For example, in 2017, the American College of Obstetricians and Gynecologists (ACOG) proposed a revised definition that included cumulative blood loss ≥ 1,000 mL or bleeding with clinical evidence of hypovolemia occurring within 24 hours of delivery.^29^ Clinical evidence of hypovolemia can be inconsistent as the signs can be non-specific and variable. Additionally, physiologic changes of pregnancy alter a woman’s tolerance for hypovolemia, making it difficult to set strict parameters for intervention based on clinical signs.^29^ These limitations suggest that it may be useful to consider additional clinically based alternative or supplementary definitions of PPH.

In this work, we propose and evaluate two primary concepts. First, we introduce a working definition of PPH based on the clinical need to intervene (cPPH). By focusing on identifying cases where treatment is deemed necessary by clinicians, this approach addresses common limitations associated with definitions based on the estimation or quantification of blood loss (EBL/QBL) and the use of non-specific clinical parameters. This cPPH definition could be used to supplement definitions based on EBL or QBL. Second, we evaluate an interpretable PPH prediction pipeline using large language models (LLMs) applied to clinical notes to extract clinically relevant features, which are then used in a predictive model.

## METHODS

### Data Source and Study Population

#### St. Luke’s University Health Network (SLUHN)

SLUHN is a non-profit, regional health network in eastern Pennsylvania and western New Jersey comprising 12 hospitals and more than 300 outpatient sites. The network’s labor and delivery services currently handle over 5,000 births annually across three facilities, serving a diverse population that includes urban, suburban, and rural communities.

#### Inclusion Criteria and required visits

We included all pregnancies within SLUHN with gestational ages ≥22 weeks of gestational age (wga), with two or more prenatal obstetric visits and at least one ultrasound between 16-24 wga. We included deliveries that occurred between January 7, 2016 and October 25, 2021.

#### Data

For all mothers in the cohort, we obtained de-identified data from the electronic medical record including structured data (i.e. patient demographics, diagnoses, labs, medications, and procedures) and clinical notes (i.e. OB Intake Interviews, ultrasounds, progress notes, and anatomy scans). Data were included beginning from the working age of conception, prioritizing gestational age derived in the first ultrasound and falling back to last menstrual period if no ultrasound was performed prior to 14 wga. Data were included up until three days prior to the mother’s arrival at the hospital for the admission which included delivery. When considering any potential deployment, predictions would be run on a daily basis after 22 weeks but would only be surfaced to clinicians when a woman arrives for a suspected delivery or induction. Data were temporally split into a training dataset (15,399 deliveries occurring between January 7, 2016, and December 31, 2019) and evaluation dataset (4,593 deliveries between January 1, 2020, and October 25, 2021).

### Natural Language Processing – Base Model

We used a large language model for three tasks, described in further detail in the following sections. We performed separate finetuning of this model for each of these tasks, which included: 1) extraction of EBL/QBL used for one definition of PPH (Table 1), 2.) direct prediction of PPH using both definitions (Fig. 1B), and 3.) extraction of clinical features for PPH prediction using both definitions (Fig. 1C). We utilized the Meta Llama-3.1-8B as our base model (https://huggingface.co/meta-llama/Llama-3.1-8B). Importantly this model has a context length of 128k, sufficient to include the full set of notes for > 99% of the included pregnancies. For cases where the total note length exceeded 128k tokens, we prioritized notes in reverse chronological order, including the most recent notes first until adding another full note would exceed the token limit.

**Figure 1.**
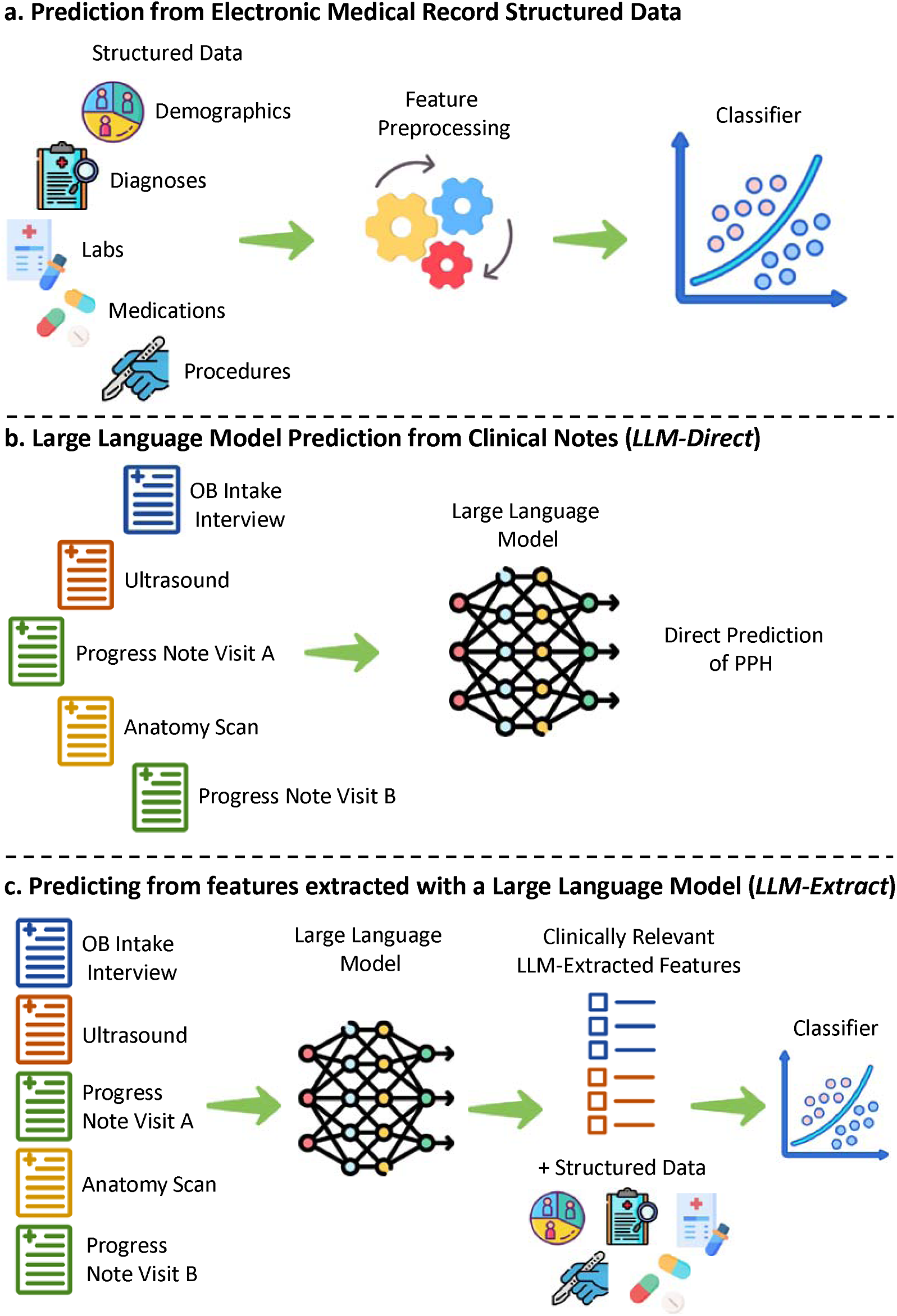
PPH Prediction Pipelines. **A.)** Predicting PPH with the structured data available in the EMR using Logistic Regression or XGBoost. **B.)** Predicting PPH using a locally pre-trained then supervised fine-tuned large language model (Llama-3.1). **C.)** Using a fine-tuned large language model to extract pre-specified clinically relevant features as input in addition to the structured data for a supervised classifier (logistic regression and XGBoost).

**Table 1.** Definitions of PPH included in the study.

| Name | Definition |
| --- | --- |
| 1. Clinical Intervention (cPPH) | At least one of the following criteria:<br>1) Transfusion of 1 or more units of red blood cells, or<br>2) Hysterectomy, or<br>3) Receipt of Bakri balloon, or<br>4) Use of three or more medications (uterotonic(s) or tranexamic acid) |
| 2. Estimated / Quantitative Blood Loss (EBL/QBL) via Clinical Notes | Extracted EBL or QBL greater than or equal to 500ml for vaginal delivery or 1000 ml for cesarean section<sup>22</sup> |

### Outcome Ascertainment and Comparison

#### Definitions of outcomes

We defined two classes of outcomes, each serving a different purpose, to assess the effects of PPH definition used for prediction (Table 1). The first class, “Clinical Intervention” (cPPH), defines PPH based on the initiation of any one of an expert-defined set of interventions based on structured data. These include transfusions, medical interventions for postpartum hemorrhage (e.g., multiple uterotonics), and procedural interventions (e.g., Bakri balloon placement, hysterectomy). The purpose of this definition is to determine when PPH is clinically meaningful in terms of necessary treatment.

The second class, “Estimated/Quantitative Blood Loss” (EBL/QBL), is based on blood loss quantification extracted through clinical notes. The study period corresponded to a transition phase in which QBL was being adopted as an institutional standard. Both EBL and QBL were used but in nearly all settings only one of the two values is available. In many cases, EBL/QBL are available in semi-structured form as part of a templated note. We first used regular expressions to extract either EBL or QBL from templated notes. For a subset of mothers flowsheet values were also available; however, these structured entries presented ascertainment challenges due to ambiguity in total EBL/QBL as well as heterogeneity throughout the health network. For example, while in some cases the total EBL/QBL was the sum of available flowsheet rows, in other cases the total EBL/QBL was the most recent flowsheet row entered. To work around this ambiguity, we finetuned an LLM to extract EBL/QBL values using examples where flowsheet values were unambiguous by identifying exact matches of either the sum or last entry within the text of a clinical note. We then validated this model by manually reviewing 250 notes for each of EBL and QBL, observing 100% inter-rater (human and LLM) agreement.

#### Outcome prediction - parameter tuning and performance evaluation

We first performed 10-fold cross validation over the training dataset to perform hyperparameter tuning of all three prediction pipelines for each of the two outcomes (Fig. 1). After all parameter tuning, we selected the top-performing models from cross-validation and applied them to the hold-out evaluation dataset. Inference on the evaluation dataset was performed a single time, and no training or additional parameter tuning was done after inference.

### PPH Prediction Pipelines

#### Structured Data Prediction

We first predicted PPH using structured data from the electronic medical record, including patient demographics, diagnoses, labs, medications, and procedures for all mothers in the cohort (Fig. 1A). Data was included from the working age of conception up until three days prior to the mother’s arrival at the hospital for delivery admission. As in Beaulieu-Jones et al.^19^ we constructed count vectors for each data attribute before performing log normalization and clipping to a maximum value of 1. This approach accounts for the signal of having multiple entries for the same attribute, while prioritizing the presence or absence of any entries and limiting the impact of outliers. For labs, we also aggregate results including the minimum value, maximum value, mean value, most recent value, count of measurements and the count of measurements outside of the reference range or abnormal values. We then trained XGBoost and logistic regression models to predict PPH based on both definitions. This was followed by a hyperparameter sweep matching the sweep performed in Beaulieu-Jones et al.^19^

#### LLM-Direct: LLM Fine-tuned to directly output Prediction of PPH

Next, we performed direct prediction of PPH (LLM-Direct) using supervised fine-tuned of an LLM, Llama-3.1 (Fig. 1B). Prediction was performed based on clinical notes including OB Intake Interviews, ultrasounds, progress notes, and anatomy scans. The LLM was fine-tuned for classification using the Hugging Face Transformers library. Parameter-Efficient Fine-Tuning (PEFT) was employed via Low-Rank Adaptation (LoRA) to optimize computational efficiency while preserving model performance. The model was trained using supervised learning on the labeled training dataset. Training was conducted using a single node of 8 A100 NVIDIA GPUs, with early stopping implemented based on validation loss (10% of training data). Hyperparameters were optimized through grid search, with the final values selected based on model performance on the validation set. Only after final parameters were selected was inference performed using the model against the hold out test data set.

#### LLM-Extract: LLM Feature Extraction for PPH Prediction

We then used the fine-tuned LLM to extract pre-specified clinically relevant features (Supplemental Table 5), which were then used along with structured data for PPH prediction (LLM-Extract, Fig. 1C). To do this, we used a knowledge distillation approach as described in Woo et al.^30^ to develop a feature extraction model using a teacher-student approach with Llama-3.1-70B-Instruct and Llama-3.1-8B-Instruct. This process both improves the model’s performance on questions where the correct answer is not available in a note and improves computational performance to make the process feasible on academic hardware. We first used Llama-3.1-70B-Instruct to generate synthetic generic feature extraction examples as in Woo et al.^30^. We provide a source document and prompt the model to provide structured outputs in JSON format containing: (1) the identified feature; (2) the feature type (e.g., categorical, numerical, temporal); (3) the extracted value; (4) the source text span; (5) a confidence score from 1-10; and (6) an explanation justifying the extraction, including relevant context from the source document. To ensure coverage, we generated examples across different feature types, including cases where features are absent or ambiguous in the source text. We then fine-tuned an instance of Llama-3.1-8B using the curated synthetic dataset with QLoRA, employing 4-bit quantization to optimize memory usage. The fine-tuning process used paged optimizers for efficient memory management between GPU and CPU resources. Based on the feature extraction task’s factual nature, we prioritize deterministic sampling strategies to maximize extraction accuracy (temperature = 0, top_p = 0.5). The explanation and source act similarly to retrieval augmented generation to minimize hallucinations^31–33^. These features were combined with the structured data described in the “Structured Data Prediction” section above and input into supervised classifiers including logistic regression and XGBoost.

#### Outcome prediction - parameter tuning and performance evaluation

We first performed 10-fold cross validation over the training dataset to perform hyperparameter tuning of all three prediction pipelines for each of the two outcomes (Fig. 1). After all parameter tuning, we selected the top-performing models from cross-validation and applied them to the hold-out evaluation dataset. Inference on the evaluation dataset was performed a single time, and no training or additional parameter tuning was done after inference.

### Data Availability Statement

Authors had primary access to all data used in this study (BKB). Access to data requires affiliation with St. Luke’s University Health Network and therefore cannot be publicly shared.

### Standard Protocol Approvals, Registrations, and Patient Consents

This study was determined to be exempt from IRB because it uses only de-identified, retrospective data and is not considered human subject’s research. Because data are retrospective, de-identified and results are published only in aggregate form, individual consent was not required for publication.

## RESULTS

### Study Population Demographics and Background

The training dataset included 15,399 deliveries occurring between January 7, 2016, and December 31, 2019. The evaluation dataset included 4,593 deliveries between January 1, 2020, and October 25, 2021. Having a temporal split between training and evaluation datasets allowed for evaluation of generalizability over time. No significant differences in the mother’s age, gestational age, prior para (live births), or delivery method were observed between the training period and evaluation period (Supplemental Table 1).

Postpartum hemorrhage was defined using two methods as previously described (Table 1, breakdown in Table 2), including a definition based on clinical intervention (cPPH) and a definition based on the validated extraction of estimated or quantitative blood loss (EBL / QBL) consistent with established definitions^24,34^. We found that there were no deliveries meeting the hysterectomy criteria without meeting other, earlier occurring criteria for cPPH. The extraction of EBL/QBL values was validated in 500 notes (250 each of EBL and QBL) without any disagreement between algorithmic and human extraction. Given that definitions of PPH based on either EBL or QBL can vary, we compared EBL/QBL between different delivery methods. Women undergoing cesarean sections averaged more than double (vaginal – mean: 345.22, median 300, c-section – mean: 753.38, median: 700) the EBL/QBL as women who gave birth vaginally (Supplemental Table 3). A larger number of mothers met the EBL/QBL-based definition than for cPPH (Table 2). There were 1,129 mothers (7.3%) in the training dataset and 336 (7.32%) in the evaluation dataset who met the EBL/QBL definition of PPH, while there were 517 mothers (3.36%) in the training dataset and 231 (5.03%) in the evaluation dataset who met the cPPH definition. Mothers who met only the cPPH definition had a slightly lower mean gestational age at delivery (261.34-262.75 days) compared to mothers who met only the EBL/QBL definition (mean 268.70-270.78 days) and mothers who met neither definition (mean 270.43-271.71 days).

**Table 2.**
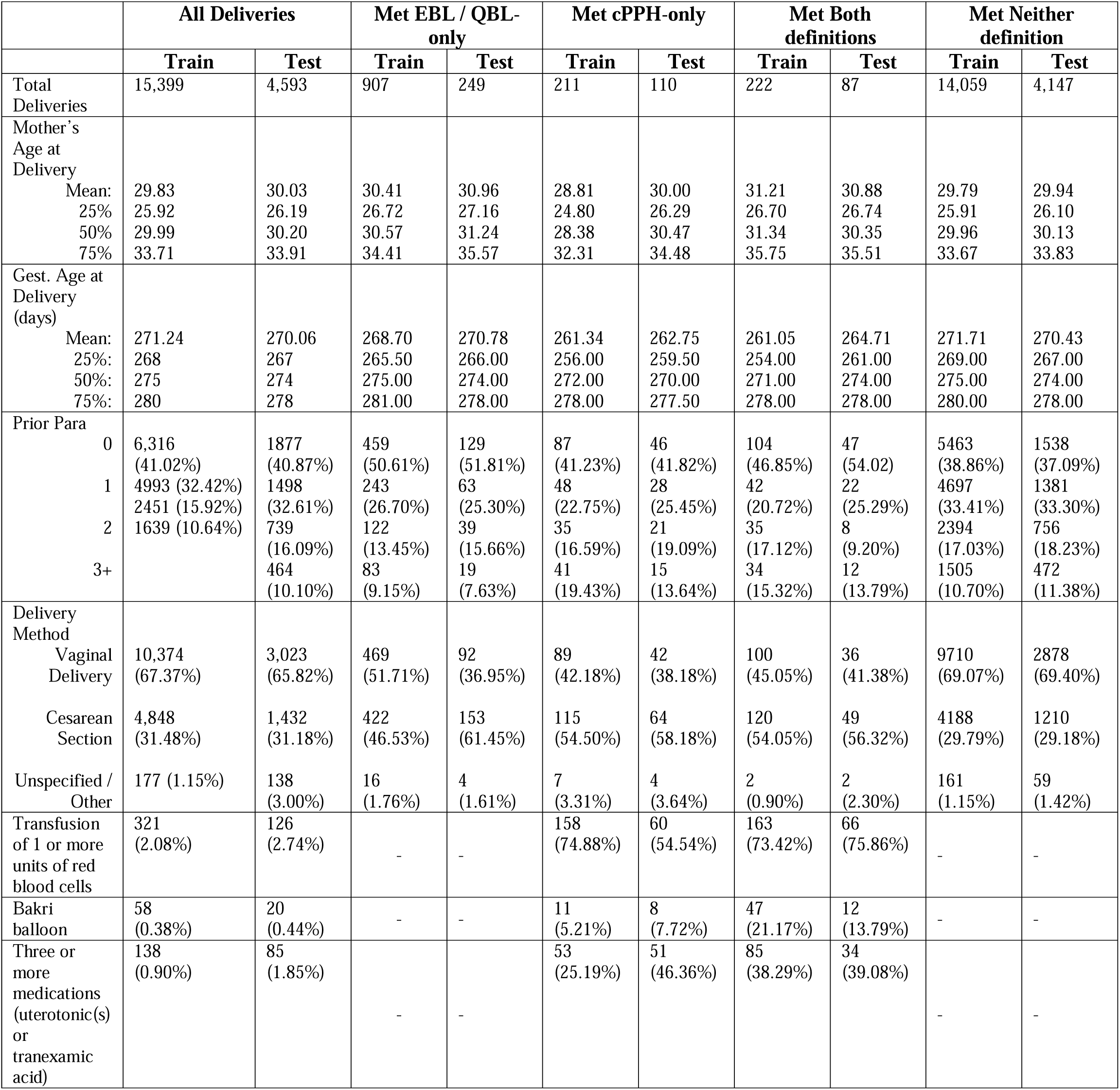
Demographic Details of Training and Evaluation Time Periods.

### Model Training and Parameter Tuning

We then trained three different predictive pipelines separately for both of the PPH definitions, using data up until three days before the mother’s delivery admission (Fig. 1, full details in Methods). The first pipeline (**Structured**) extracts and performs feature preprocessing from the structured data (demographics, diagnoses, labs, medications, and procedures) in the EMR (Fig. 1A). The second pipeline (**LLM-Direct**) finetunes a large language model, Llama 3.1, to predict PPH from the concatenated clinical notes prior to the delivery admission (Fig. 1B). Using a LLM to directly predict PPH can be difficult to validate or interpret using current methods. In contrast, training a LLM to extract features allows for performance validation by enabling manual chart review for a small subset of mothers to verify accurate feature extraction. Therefore, the third pipeline (**LLM-Extract**) finetunes Llama-3.1 to extract structured (e.g., binary or numeric) clinical features about the symptoms, medical and family histories, measurements from imaging reports and other expert specified features which may be relevant to PPH (Features listed in Supplemental Table 4). These LLM-extracted features are combined with existing structured data (as in the Structured pipeline), which are then used for PPH prediction with logistic regression and XGBoost models (Fig. 1C).

During training of the models, we performed parameter selection and tuning using 10-fold cross validation of the training data (Table 3). Performance of the two models was higher in the cPPH definition for all pipelines, which may be in part due to the variation in the number of mothers matching each definition (Table 2). For both PPH definitions, we observed highest performance using the LLM for direct PPH prediction (LLM-Direct), with a Mean AUROC 0.844 (0.818-0.869) for cPPH and 0.792 (0.764-0.830) for EBL/QBL. The XGBoost classifier for LLM-Extract did not perform as well as the direct LLM-Direct for either the cPPH definition (Mean AUROC = 0.814) or EBL/QBL definition (Mean AUROC = 0.730), but slightly outperformed logistic regression LLM-Extract (cPPH Mean AUROC = 0.794; EBL/QBL Mean AUROC = 0.721).

**Table 3.** Ten-fold cross validation performance of classifiers on training data (included deliveries between 1/1/2015 and 12/31/2019, mean (95% CI)).

|  |  | AUROC |  |
| --- | --- | --- | --- |
|  |  | cPPH | EBL / QBL |
| Structured Data Only | Logistic regression | 0.708<br>(0.683-0.731) | 0.656<br>(0.637-0.667) |
|  | XGBoost | 0.730<br>(0.711-0.746) | 0.682<br>(0.668-0.694) |
| LLM-Direct | LLM | 0.844<br>(0.818-0.869) | 0.792<br>(0.764-0.830) |
| LLM-Extract | Logistic Regression | 0.794<br>(0.770 -0.818) | 0.721<br>(0.704-0.733) |
|  | XGBoost | 0.814<br>(0.791-0.831) | 0.730<br>(0.718-0.748) |

### Model Evaluation

We then compared the performance of the direct LLM classifier (LLM-Direct) to both XGBoost and Logistic Regression models using the LLM extracted features (LLM-Extract) on the test set. Again, LLM- Direct (AUROCs of 0.79-0.80 across) slightly outperformed both logistic regression and XGBoost LLM- Extract models (AUROCs of 0.76-0.78) for both PPH definitions (Fig. 2). While the LLM-Extract models approached performance of LLM-Direct, LLM-Extract clearly outperformed models that used only structured data (AUROCs 0.65-0.71). Between the models using only structured data, performance was slightly higher with the EBL/QBL definition (Logistic Regression AUROC = 0.69; XGBoost AUROC = 0.71) compared to the cPPH definition (Logistic Regression AUROC = 0.66; XGBoost AUROC = 0.65). For the LLM-Direct and LLM-Extract models, performance was similar between EBL/QBL and cPPH definitions.

**Figure 2.**
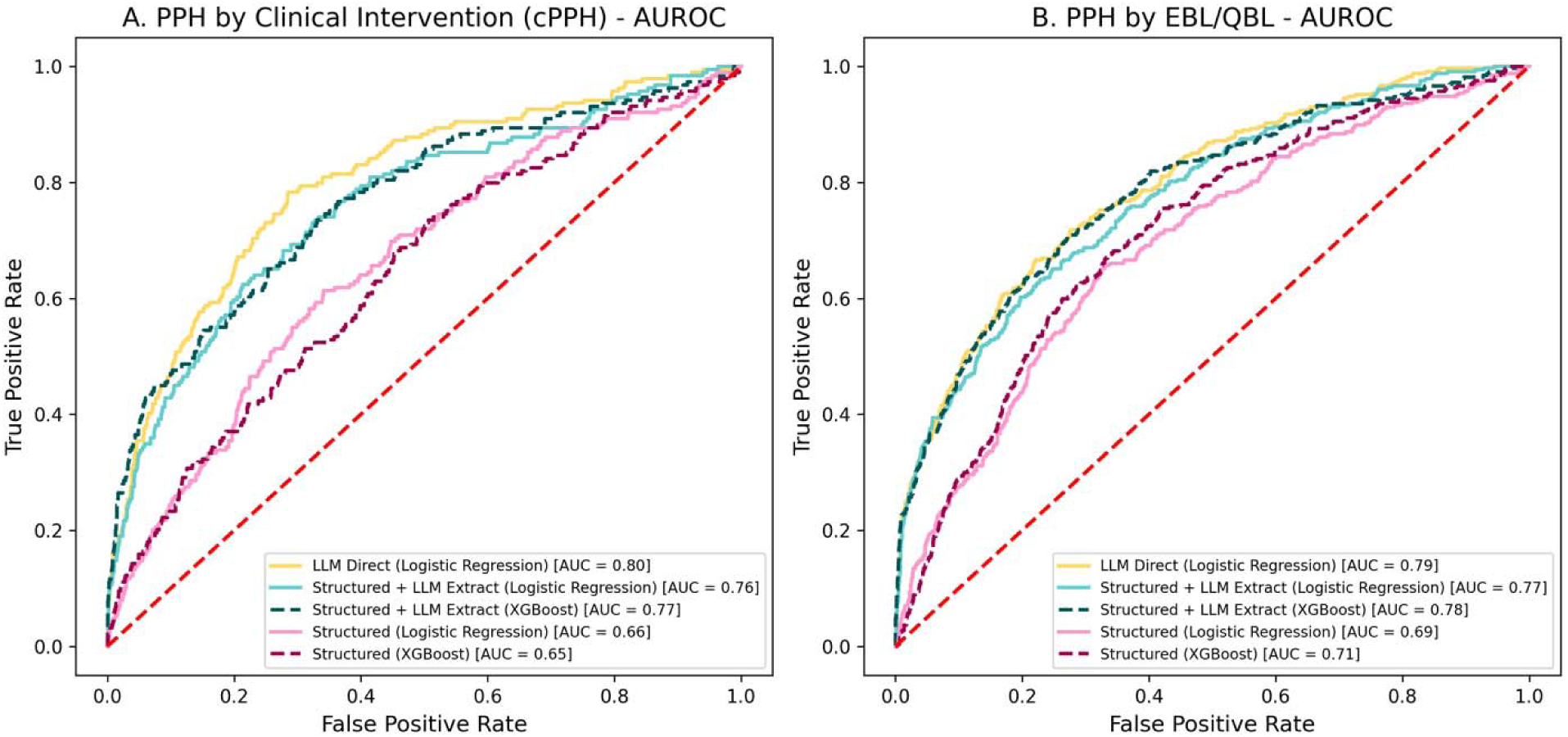
Test set performance for LLM and LLM Extracted Models.

### Feature Importance

There are several reasons why a direct LLM classifier cannot be directly deployed in an EMR, including a lack of interpretability. Additionally, there is a possibility that such a classifier could train clinicians to write notes in a manner which provides the output they desire. Given these limitations, we wanted to further interrogate the LLM-Extract models to identify a subset of features which are useful to predict PPH. If successful, this could enable prospective screening or simple calculators to help identify women at increased risk of PPH. We assessed feature significance in the logistic regression model (Structured + LLM Extract, using the EBL/QBL definition of PPH) and identified 47 significant features (Table 4). For example, multiple gestation (adj. p-val = 1.62E-21; OR 1.28 [1.22-1.35]) was a note-extracted feature found in 4.1% (638/15,399) of the deliveries from the training period. Previous cesarean (adj. p-val = 3.16E-08; OR 1.21 [1.13-1.30]; frequency 17.8%) and prior PPH (adj. p-val<0.001; OR 1.15 [1.06-1.24]) were also note-extracted features that are well-known risk factors for PPH. We also identified significant features from complete blood counts (CBCs) including abnormal platelet count, high leukocytes, and high immature granulocytes. Maximum platelet count (adj. p-value = 0.000883; OR 0.79 [0.69-0.91]) corresponds with previous work that suggests high pre-birth platelet count in the third trimester may be associated with reduced risk of PPH.^35^

**Table 4.**
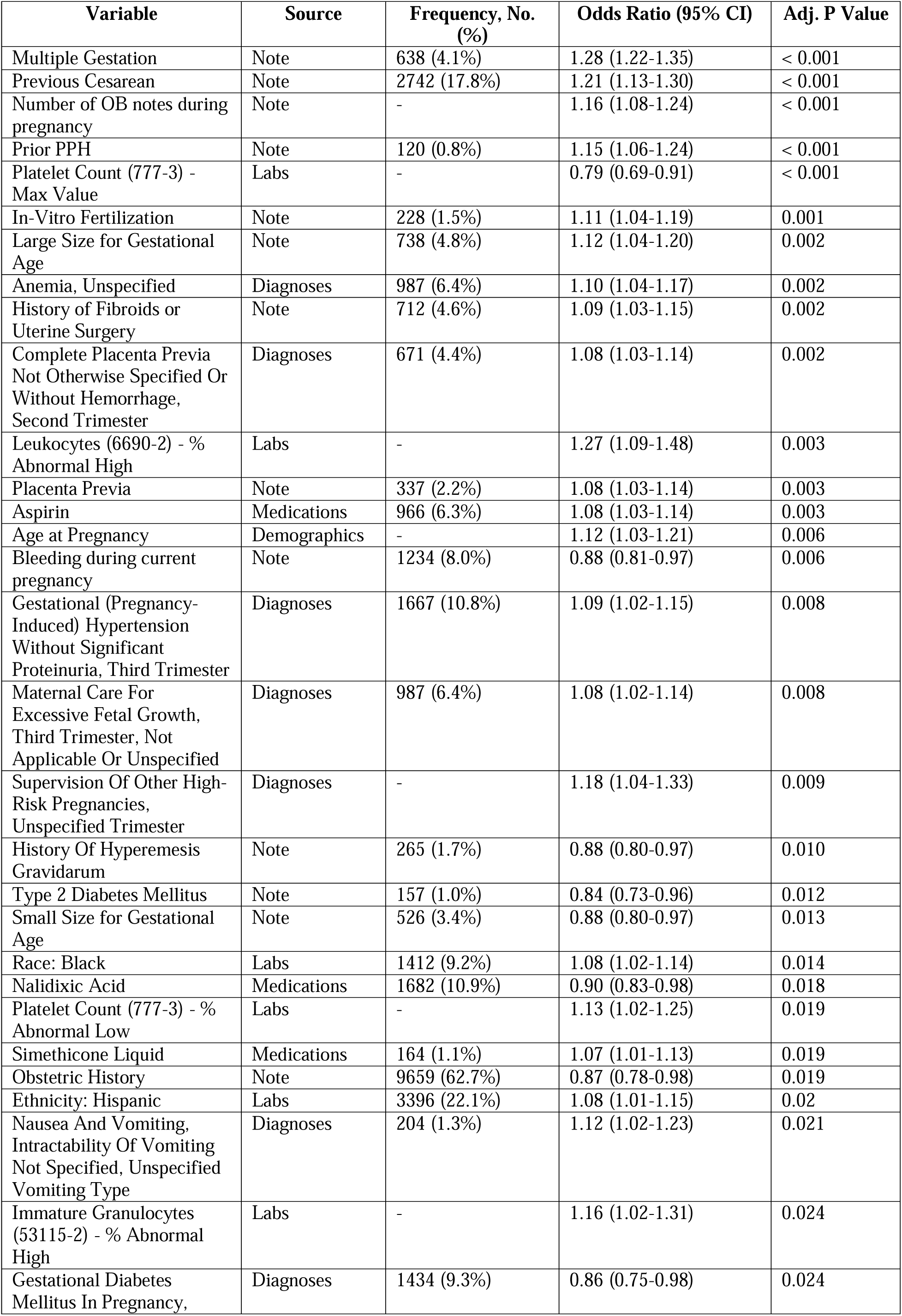
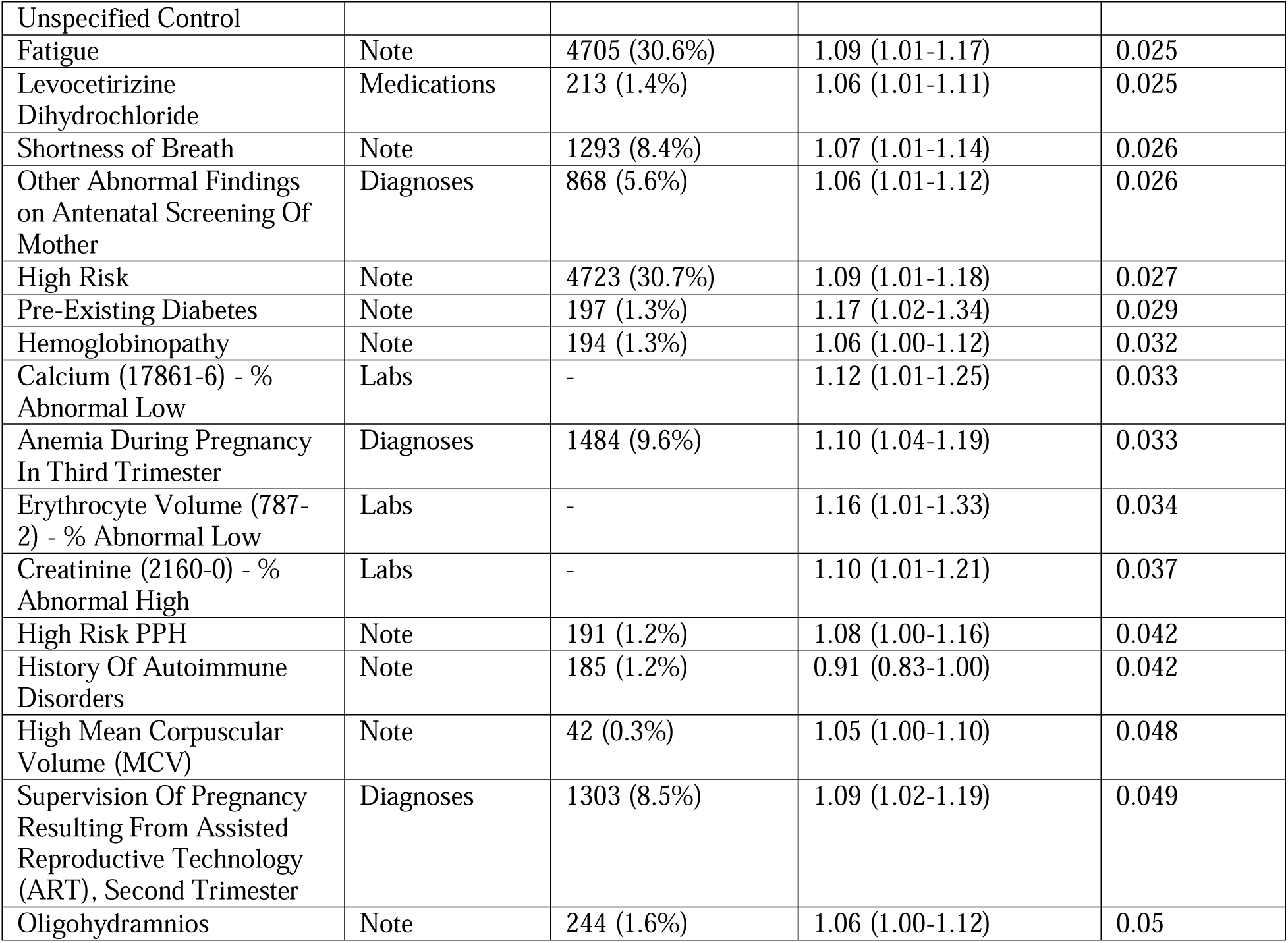
Significant (adjusted p-value < 0.05) features and their frequencies among deliveries during the training period. Prompts used to extract each variable’s name are available in Supplemental Table 5.

## Discussion

In this proof-of-concept study, we find that PPH prediction benefits from the addition of information not contained in the structured portions of the EMR. Fine-tuned LLMs outperform predictive models built on structured data. Importantly, all models do not include any data from the delivery and the predictions are designed to be used immediately when labor is suspected, or induction is attempted. Models which use data from the admission may be susceptible to hindsight bias where information is assumed to be available but was in fact captured after the fact. For example, if a vaginal delivery is attempted but complications necessitate a transition to cesarean section, the type of delivery is a challenging feature to use in a prediction model.

There are often limitations to clinical risk prediction using large language models, including: 1) the lag between the time an event occurs and the time a clinical note is finalized, 2) the lack of either interpretability or explainability of LLM-based predictions, and 3) the likelihood that the deployment of a risk model based solely on clinical notes would “train” physicians and other clinicians to write notes in ways to encourage the LLM’s predictions to match their intuition. For example, if a physician believes a patient is at high risk of PPH and would benefit from potential intervention (such as closer monitoring or delivery at a specific hospital within the network), the physician might learn to write notes in a way to ensure the patient receives that intervention.^36^ To address these challenges in direct LLM prediction and to provide a better understanding of the risk factors of PPH, we introduce a new feature-based approach (LLM-Extract) for PPH prediction. LLM-Extract leverages the strengths of LLMs while performing prediction with simpler, more interpretable models. This extract-then-predict approach performs almost as well as direct LLM prediction and clearly outperforms prediction using only structured data. Using this approach, we can identify a smaller set of verifiable features which leads to more interpretable models. Our feature significance analysis identified well-established risk factors for PPH such as multiple gestation, previous cesarean, and prior PPH. This method also has the potential to identify additional risk factors that could be investigated in further studies. Replication of this work in additional populations is necessary for rigorous evaluation of features associated with PPH risk. We believe our pipeline has the potential to enable these evaluations at scale.

LLM-Extract presents several new promising opportunities for follow-up studies. First, running this pipeline on a larger population would allow for both validation of predictive performance as well as interpretability analyses. Within this work, interpretation is limited due to sample size at several levels: the cohort sample size (19,992 between both training and evaluation sets), the relatively low incidence of PPH, and the frequency of individual symptoms the model uses to predict. For example, macrosomia has been previously linked to PPH^37,38^ but it is only present in four PPH deliveries in the current training population. Given these challenges around sample size, we believe any interpretability analysis is premature. There are also limitations in model calibration at the current sample size. The predicted probabilities across all three pipelines are less than 0.2 for most mothers. The model is well calibrated at low risks where there is a large sample size, but there is substantial variation at high predicted risk probabilities. While the average predicted risk within those who have PPH is more than five times that of those who do not have PPH, the lack of stability in calibration indicates the need for validation of all three pipelines in populations with large sample sizes with PPH.

We also explore two definitions of PPH, including (i) a more traditional EBL/QBL-based definition extracted from notes and (ii) a new clinical intervention-based (cPPH) definition using structured data. Identification of necessary treatment for cPPH can occur during or after the birth and is not reliant on real time recognition during delivery. Identifying the population who meet only one of these two definitions (cPPH or EBL/QBL) can be informative of whether true cases of PPH are missed under either of these definitions, as well as of the concordance of these definitions with one another.

While in the current study we focus primarily on the ability of LLM-Extract to identify a subset of key features for prediction models, we hope to validate this approach in larger cohorts to enable the development of deployable risk models or guided survey-based tools. Successful deployment of any such tool is reliant on appropriate and effective intervention. Current actions based on model output are generally geared at management of PPH as opposed to prevention. Notably, our approach predicts PPH prior to the delivery and does not require data from the delivery admission, including previously reported risk factors such as details of the labor course, duration, and delivery type. An accurate risk model with the ability to predict PPH prior to delivery could enable more effective assessment as well as clinical studies of potential preventative interventions.

## ACKNOWLEDGEMENTS

We would like to acknowledge St. Luke’s logistical and technical support provided by Dr. James Balshi, Dr. Charles Sonday (Network Medical Director of Informatics), and Dr. Aldo Carmona (Senior Vice President for Medical Integration).

## AUTHOR CONTRIBUTIONS

| Name | Conception or design of the work | Acquisition/ Preprocessing of data | Analysis / Interpretation of Data | Manuscript draft | Revision |
| --- | --- | --- | --- | --- | --- |
| Brett Beaulieu-Jones | x | x | x | x | x |
| Tyler Gifford | x | x | x | x | x |
| Geena Woo |  |  | x | x | x |
| Israel Zighelboim | x | x | x | x | x |
| Joseph Bell | x |  | x | x | x |
| Jorge E. Tolosa | x | x | x | x | x |
| Hannah Milthorpe |  |  | x | x | x |
| Emily Alsentzer |  |  | x | x | x |
| Ryan Longman |  |  |  | x | x |

## Notes

### Competing Interest Statement

The authors have declared no competing interest.

### Funding Statement

This work was funded in part by the National Institutes of Health, specifically grant number R00NS114850 to BKB.

### Author Declarations

The ethics committee/IRB of St. Lukes University Hospital Network waived ethical approval for this work. Because data are retrospective and de-identified and results are published only in aggregate form individual consent was not required for publication.

## REFERENCES

1. Say L, Chou D, Gemmill A, et al. Global causes of maternal death: a WHO systematic analysis. Lancet Glob Health. 2014;2(6):e323–33.

2. Bingham D, Melsop K, Main E. CMQCC obstetric hemorrhage hospital level implementation guide. The California Maternal Quality Care Collaborative (CMQCC). Published online 2010.

3. Postpartum Hemorrhage (PPH). AWHONN. January 5, 2020. Accessed October 18, 2024. https://www.awhonn.org/postpartum-hemorrhage-pph/

4. Dmission D a. Risk Assessment Tables. https://www.acog.org/-/media/project/acog/acogorg/files/forms/districts/smi-ob-hemorrhage-bundle-risk-assessment-ld-admin-intrapartum.pdf

5. Dilla AJ, Waters JH, Yazer MH. Clinical validation of risk stratification criteria for peripartum hemorrhage. Obstet Gynecol. 2013;122(1):120–126.

6. Kawakita T, Mokhtari N, Huang JC, Landy HJ. Evaluation of risk-assessment tools for severe postpartum hemorrhage in women undergoing cesarean delivery. Obstet Gynecol. 2019;134(6):1308–1316.

7. Zheutlin AB, Vieira L, Shewcraft RA, et al. Improving postpartum hemorrhage risk prediction using longitudinal electronic medical records. J Am Med Inform Assoc. 2022;29(2):296–305.

8. Antepartum Screening and Rapid Response Reduce Postpartum Hemorrhage by 40 Percent. EpicShare. Accessed February 18, 2025. https://www.epicshare.org/share-and-learn/antepartum-screening-and-rapid-response-reduce-postpartum-hemorrhage-by-40-percent

9. Neary C, Naheed S, McLernon DJ, Black M. Predicting risk of postpartum haemorrhage: a systematic review. BJOG. 2021;128(1):46–53.

10. Koopmans CM, van der Tuuk K, Groen H, et al. Prediction of postpartum hemorrhage in women with gestational hypertension or mild preeclampsia at term. Acta Obstet Gynecol Scand. 2014;93(4):399–407.

11. Venkatesh KK, Strauss RA, Grotegut CA, et al. Machine learning and statistical models to predict postpartum hemorrhage. Obstet Gynecol. 2020;135(4):935–944.

12. Liu J, Wang C, Yan R, et al. Machine learning-based prediction of postpartum hemorrhage after vaginal delivery: combining bleeding high risk factors and uterine contraction curve. Arch Gynecol Obstet. 2022;306(4):1015–1025.

13. Aghajanian S, Jafarabady K, Abbasi M, et al. Prediction of post-delivery hemoglobin levels with machine learning algorithms. Sci Rep. 2024;14(1):13953.

14. Westcott JM, Hughes F, Liu W, Grivainis M, Hoskins I, Fenyo D. Prediction of maternal hemorrhage using machine learning: Retrospective cohort study. J Med Internet Res. 2022;24(7):e34108.

15. Agrawal M, Hegselmann S, Lang H, Kim Y, Sontag D. Large language models are few-shot clinical information extractors. arXiv [csCL]. Published online May 25, 2022. http://arxiv.org/abs/2205.12689

16. Wei J, Bosma M, Zhao VY, et al. Finetuned Language Models Are Zero-Shot Learners. arXiv [csCL]. Published online September 3, 2021. http://arxiv.org/abs/2109.01652

17. Thompson WE, Vidmar DM, De Freitas JK, et al. Large Language Models with Retrieval-Augmented Generation for Zero-Shot Disease Phenotyping. arXiv [csAI]. Published online December 11, 2023. http://arxiv.org/abs/2312.06457

18. Alsentzer E, Rasmussen MJ, Fontoura R, et al. Zero-shot interpretable phenotyping of postpartum hemorrhage using large language models. NPJ Digit Med. 2023;6(1):212.

19. Beaulieu-Jones BK, Villamar MF, Scordis P, et al. Predicting seizure recurrence after an initial seizure-like episode from routine clinical notes using large language models: a retrospective cohort study. Lancet Digit Health. 2023;5(12):e882–e894.

20. Jiang LY, Liu XC, Nejatian NP, et al. Health system-scale language models are all-purpose prediction engines. Nature. 2023;619(7969):357–362.

21. McInerney D, Young G, van de Meent JW, Wallace B. CHiLL: Zero-shot custom interpretable feature extraction from clinical notes with large language models. In: Findings of the Association for Computational Linguistics: EMNLP 2023. Association for Computational Linguistics; 2023:8477–8494.

22. Rath WH. Postpartum hemorrhage--update on problems of definitions and diagnosis. Acta Obstet Gynecol Scand. 2011;90(5):421–428.

23. Who IV. Prevention and management of postpartum haemorrhage. Report of a technical working group. Geneva 3-6 July 1989. World Health Organization/Maternal and Child Health. Published online 1990.

24. OB Hemorrhage Toolkit V3.0. Accessed October 24, 2024. https://www.cmqcc.org/resources-tool-kits/toolkits/ob-hemorrhage-toolkit

25. Wormer KC, Jamil RT, Bryant SB. Postpartum hemorrhage. In: StatPearls. StatPearls Publishing; 2024.

26. Quantitative Blood Loss in Obstetric Hemorrhage. Accessed November 14, 2024. https://www.acog.org/clinical/clinical-guidance/committee-opinion/articles/2019/12/quantitative-blood-loss-in-obstetric-hemorrhage

27. Diaz V, Abalos E, Carroli G. Methods for blood loss estimation after vaginal birth. Cochrane Database Syst Rev. 2018;9(9):CD010980.

28. Zhang WH, Deneux-Tharaux C, Brocklehurst P, et al. Effect of a collector bag for measurement of postpartum blood loss after vaginal delivery: cluster randomised trial in 13 European countries. BMJ. 2010;340(feb01 1):c293.

29. Borovac-Pinheiro A, Pacagnella RC, Cecatti JG, et al. Postpartum hemorrhage: new insights for definition and diagnosis. Am J Obstet Gynecol. 2018;219(2):162–168.

30. Woo EG, Burkhart MC, Alsentzer E, Beaulieu-Jones BK. Synthetic data distillation enables the extraction of clinical information at scale. medRxiv. Published online September 28, 2024:2024.09. 27.24314517. doi:10.1101/2024.09.27.24314517

31. Niu C, Wu Y, Zhu J, et al. RAGTruth: A hallucination corpus for developing trustworthy retrieval-augmented language models. arXiv [csCL]. Published online December 30, 2023. http://arxiv.org/abs/2401.00396

32. Béchard P, Ayala OM. Reducing hallucination in structured outputs via Retrieval-Augmented Generation. arXiv [csLG]. Published online April 11, 2024. http://arxiv.org/abs/2404.08189

33. Tonmoy SMTI, Zaman SMM, Jain V, et al. A comprehensive survey of hallucination mitigation techniques in Large Language Models. arXiv [csCL]. Published online January 2, 2024. https://www.amanchadha.com/research/2401.01313.pdf

34. Rath WH. Postpartum hemorrhage--update on problems of definitions and diagnosis: Postpartum hemorrhage - definitions and diagnosis. Acta Obstet Gynecol Scand. 2011;90(5):421–428.

35. Robinson MR, Patxot M, Stojanov M, Blum S, Baud D. Postpartum hemorrhage risk is driven by changes in blood composition through pregnancy. Sci Rep. 2021;11(1):19238.

36. Beaulieu-Jones BK, Yuan W, Brat GA, et al. Machine learning for patient risk stratification: standing on, or looking over, the shoulders of clinicians? NPJ Digit Med. 2021;4(1):62.

37. Beta J, Khan N, Fiolna M, Khalil A, Ramadan G, Akolekar R. Maternal and neonatal complications of fetal macrosomia: cohort study. Ultrasound Obstet Gynecol. 2019;54(3):319–325.

38. Alsammani MA, Ahmed SR. Fetal and maternal outcomes in pregnancies complicated with fetal macrosomia. N Am J Med Sci. 2012;4(6):283–286.

