## Supplemental Tables 1-4 for "Leveraging Large Language Models to Develop an Interpretable Prediction Model for Postpartum Hemorrhage Prior to the Onset of Labor"

**Supplemental Table 1. “Clinical Intervention outcomes” per rule.** Number of deliveries during each study period matching each intervention rule. (No women had a hysterectomy without meeting other criteria)

|  | Training Period  (01/07/2016 -12/31/2019) | Evaluation period  (01/01/2020-10/25/2021) |
| --- | --- | --- |
| Total Deliveries | 15,399 | 4,593 |
| Total PPH by Clinical Intervention Rules |  |  |
| Transfusion | 321 (2.08%) | 126 (2.74%) |
| Bakri Balloon | 58 (0.38%) | 20 (0.44%) |
| Medications | 138 (0.90%) | 85 (1.85%) |
| Combined | 433 (2.81%) | 197 (4.29%) |

**Supplemental Table** **2**. Estimated and Quantitative Blood Loss for each delivery.

|  | Training | Evaluation |
| --- | --- | --- |
| Total Births | 15,399 | 4,593 |
| Mean Blood Loss | 519.19 | 444.51 |
| 25% | 300 | 200 |
| 50% | 449.5 | 350 |
| 75% | 700 | 700 |

**Supplemental Table** 3. Estimated EBL/QBL stratified by delivery method.

|  | Vaginal Delivery (ml) | C-Section (ml) |
| --- | --- | --- |
| Mean | 345.22 | 753.38 |
| 25% | 125 | 600 |
| 50% | 300 | 700 |
| 75% | 350 | 800 |
| Max | 3527 | 5500 |

**Supplemental Table 4. Comparison between Label Source.**

|  | **Training** | **Test** |
| --- | --- | --- |
| EBL/QBL only | 834 | 224 |
| Structured only | 222 | 119 |
| Both Definitions | 295 | 112 |

**Supplemental Table 5**. Features for LLM Extraction.

| **Key** | **Question** |
| --- | --- |
| high_risk | Do the notes state that this is a high-risk pregnancy? |
| high_risk_bleeding | Do the notes state any evidence of increased risk of bleeding? |
| high_risk_pph | Do the notes state any evidence of increased risk of postpartum hemorrhage? |
| high_risk_adv | Do the notes state any increased risk of adverse outcomes during delivery? |
| small_size | Do the notes state the baby is small for gestational age? |
| large_size | Do the notes state the baby is large for gestational age? |
| nausea | Has the patient experienced nausea? |
| vomiting | Has the patient experienced vomiting? |
| asthma | Does the patient have asthma? |
| sob | Has the patient experienced any shortness of breath? |
| constipation | Has the patient experienced constipation? |
| headaches | Has the patient experienced headaches? |
| fatigue | Has the patient experienced fatigue? |
| cramping | Has the patient experienced cramping? |
| spotting | Has the patient had any spotting or any other bleeding? |
| cravings | Has the patient had any cravings? |
| GDM | Does the patient have a history of gestational diabetes mellitus (GDM)? |
| PCOS | Does the patient have a history of PCOS? |
| t2dm | Does the patient have type 2 diabetes? |
| endometriosis | Does the patient have endometriosis? |
| fam_t2dm | Does the patient have a first degree relative with type 2 diabetes? |
| bloating | Has the patient experienced bloating? |
| lightheadedness | Has the patient experienced lightheadedness? |
| dizziness | Has the patient experienced dizziness? |
| BMI_35 | Does the patient have a BMI over 35? |
| BMI_30 | Does the patient have a BMI over 30? |
| weight_loss | Has the patient had abnormal weight loss? |
| weight_gain | Has the patient had abnormal weight gain? |
| obesity | Is the patient obese? |
| underweight | Is the patient underweight? |
| weight_change | Do the notes mention concern about appropriate weight gain during pregnancy? |
| htn | Does the patient have a history of hypertension or HTN? |
| fetus_gender | Is the fetus a male? |
| chronic_htn | Does the patient have a history of chronic or ongoing hypertension? |
| ckd | Does the patient have a history of chronic kidney disease or CKD? |
| t1d | Does the patient have a history of type 1 diabetes? |
| infection | Has the patient had any noted infections? |
| infection_abs | Has the patient needed to take any antibiotics during pregnancy? |
| mrsa | Does the patient have a history of MRSA? |
| herpes | Does the patient have a history of herpes? |
| smoking | Does the patient smoke or use tobacco? |
| alcohol | Does the patient drink alcohol? |
| illicit_substances | Does the patient use any illicit or illegal substances? |
| metformin | Does the patient take metformin? (Answer yes or no, or NA if not available in the note) |
| bleeding | Has the patient had any issues with bleeding? |
| uterine_bleeding | Has the patient experienced any vaginal or uterine bleeding? |
| fertility | Did the patient receive fertility treatment? |
| ivf | Did the patient undergo a frozen embryo transfer (FET)? |
| iui | Was the fetus conceived using Intrauterine insemination? |
| cervical_dilation | Was the cervix dilated? |
| cervical_shortening | Was any cervical shortening observed? |
| cervical_polyps | Were any cervical polyps observed? |
| cervical_polyp_removal | Were any cervical polyps removed? |
| cervical_abnormalities | Were any cervical abnormalities noted? |
| efw_normal | Was the estimated fetal weight abnormally high? |
| uterine_polyps | Is there a history of uterine polyps? |
| hysteroscopy | Has the patient undergone hysteroscopy? |
| salpingectomy | Has the patient undergone salpingectomy? |
| congenital_uterine_anomoalies | Were any congenital uterine anomalies observed? |
| congenital_septate | Was the uterus observed to be septate? |
| congenital_bicornuate | Was the uterus observed to be bicornuate? |
| congenital_unicornuate | Was the uterus observed to be unicornuate? |
| congential_uterus_didelphys | Was uterus didelphys observed? |
| fibroids | Did the patient currently have any fibroids? |
| dilation_and_curettage | Does the patient have a history of dilation and curettage? |
| macrosomia | Has the patient experienced macrosomia? |
| magnesium_sulfate | Has the patient taken magnesium sulfate during the pregnancy? |
| prior_pph | Has the patient experienced any prior postpartum hemorrhage or PPH? |
| prior_blood_loss | If the patient has previously delivered another baby, was there any excessive blood loss? |
| prior_c_section | Has the patient undergone a prior cesarean birth or c-section? |
| placenta_abruption | Do the notes mention placenta abruption or any concern about placenta abruption? |
| placenta_previa | Is placenta previa or its abbreviation mentioned in the patient’s clinical notes? |
| placenta_accreta | Do the notes mention placenta accreta or any concern about placenta accreta? |
| pas | Do the notes mention placenta accreta spectrum (PAS)? |
| coagulopathy | Do the notes mention any history of coagulopathy or other bleeding disorders? |
| platelet | Do the notes mention any history of platelet dysfunction? |
| platelet_counts | Do the notes mention any history of abnormal platelet counts? |
| fibrinogen_threshold | Do the notes mention any abnormal fibrinogen levels? |
| thrombocytopenia | Is there any mention of thrombocytopenia (low platelet count) or platelet count below 150,000/¬µL? |
| thrombocytopenia_level | Do the notes mention any low platelet counts? |
| prolonged_pt | Do the notes mention any high prothrombin time (PT) or activated partial thromboplastin times (aPTT)? |
| amniotic_fluid_embolism | Do the notes mention any issues with amniotic fluid embolisms? |
| hemolysis | Do the notes mention hemolysis? |
| liver_enzymes | Do the notes mention any elevated liver enzymes? |
| VonWIllebrand | Do the notes mention any history of Von Willebrand disease, hemopphilia or other similar bleeding disorders? |
| Asian | Is the patient Asian? |
| AfricanAmerican | Is the patient African American? |
| Hispanic | Is the patient Hispanic? |
| obstetric_history | Does the patient have a prior obstetric history? |
| obstetric_history_ptb | Does the patient have a history of preterm delivery? |
| dx_bleeding | Do the notes mention the patient having a history of any bleeding issues? |
| dx_depression | Do the notes mention the patient having a history or diagnosis of Depression? |
| dx_pysch | Do the notes mention the patient having a history or diagnosis of any psychiatric conditions? |
| dx_subchorionic | Do the notes mention the patient having a history or diagnosis of a subchorionic hemorrhage? |
| prenatal_vitamin | Do the notes indicate a prenatal vitamin is being taken? |
| vaginal_bleeding | Do the notes mention any vaginal bleeding? |
| vaginal_fluid | Do the notes mention any vaginal fluid leakage? |
| decreased_fetal_activity | Do the notes mention any decreased fetal activity? |
| abnormal_hr | Do the notes mention the fetus has an abnormal heart rate? |
| high_hr | Do the notes mention the fetus has a high heart rate? |
| low_hr | Do the notes mention the fetus has a low heart rate? |
| fetal_mvmnt | Do the notes mention any concerns about fetal movement? |
| abdominal_pain | Do the notes mention the patient having any abdominal pain? |
| headache_persistent | Do the notes mention the patient having a headache that doesn't go away with acetaminophen? |
| vision_changes | Do the notes mention the patient having any vision changes? |
| diarrhea | Do the notes mention the patient having diarrhea? |
| early_contractions | Do the notes mention the patient having early contractions? |
| anxiety | Do the notes mention a diagnosis of anxiety? |
| maternal_age | What was the maternal age at the start of pregnancy according to the notes? |
| gravida | What is the gravida (number of total pregnancies) as stated in the notes? |
| parity | What is the parity (number of live births) as mentioned in the clinical notes? |
| pre_eclampsia | Do the notes mention pre-eclampsia or its abbreviations like PEC or PET? |
| gestational_hypertension | Is there a mention of gestational hypertension (GHTN) in the patient history? |
| gestational_diabetes | Do the notes indicate that the patient had gestational diabetes (GDM)? |
| previous_cesarean | Do the notes mention any history of previous cesarean section (C-section)? |
| previous_pph | Is there documentation in the notes indicating a history of postpartum hemorrhage (PPH)? |
| multiple_gestation | Do the clinical notes reference a multiple gestation (twins, triplets, etc.) for this pregnancy? |
| placental_abruption | Do the notes refer to a history or diagnosis of placental abruption (abruption placentae)? |
| anemia | Do the notes mention anemia, hemoglobin less than 10g/dL, or the abbreviation "Hb"? |
| ethnicity | What ethnicity is stated in the notes? Specifically mention if the patient is Asian or Hispanic. |
| pre_pregnancy_hypertension | Do the notes indicate that the patient had chronic or pre-pregnancy hypertension? |
| HELLP_syndrome | Is HELLP syndrome (Hemolysis, Elevated Liver enzymes, and Low Platelet count) mentioned in the notes? |
| uterine_fibroids | Do the notes mention the presence of uterine fibroids (leiomyomas)? |
| smoking_during | Do the notes state that the patient smoked during pregnancy? |
| smoking_hx | Has the patient ever been a smoker? |
| pre_pregnancy_bmi | What is the pre-pregnancy BMI as recorded in the notes, especially if it is over 30? |
| anticoagulant_use | Do the clinical notes reference any use of anticoagulants like heparin, warfarin, or aspirin? |
| antiplatelet_use | Is there any mention of the patient being on antiplatelet therapy, such as aspirin or clopidogrel, during pregnancy? |
| HIV_status | Do the notes indicate that the patient has HIV or is receiving antiretroviral therapy? |
| pre_existing_diabetes | Is there any mention of pre-existing type 1 or type 2 diabetes before this pregnancy? |
| renal_disease | Do the clinical notes mention chronic kidney disease (CKD) or any renal impairment before or during pregnancy? |
| hepatic_disease | Is there any reference to pre-existing liver disease or conditions like hepatitis B or C in the notes? |
| intrauterine_growth_restriction | Do the notes mention intrauterine growth restriction (IUGR) or suggest fetal growth below the 10th percentile? |
| history_of_thromboembolism | Is there any history of venous thromboembolism (VTE) or deep vein thrombosis (DVT) mentioned in the patient‚Äôs history? |
| antidepressant_use | Do the clinical notes indicate that the patient was taking antidepressants during pregnancy? |
| chronic_anemia | Do the notes mention chronic anemia or a consistently low hemoglobin level prior to or during pregnancy? |
| pulmonary_disease | Do the notes indicate any history of chronic pulmonary disease such as asthma or chronic obstructive pulmonary disease (COPD)? |
| cardiac_disease | Is there any mention of pre-existing cardiac disease or conditions such as heart failure or arrhythmias in the notes? |
| hemoglobinopathy | Do the clinical notes mention any hemoglobinopathies such as sickle cell disease or thalassemia? |
| immune_conditions | Do the notes mention any autoimmune or immunosuppressive conditions in the patient‚Äôs history (e.g., lupus, rheumatoid arthritis)? |
| preterm_labor_history | Do the notes mention a history of preterm labor (before 37 weeks gestation) in prior pregnancies? |
| chronic_hypertension | Is there documentation in the notes of chronic hypertension prior to or during pregnancy? |
| breech_presentation | Do the clinical notes mention a breech or abnormal fetal presentation earlier in pregnancy? |
| preexisting_renal_disease | Is there mention of pre-existing renal disease (e.g., chronic kidney disease) in the notes? |
| polyhydramnios | Do the notes mention polyhydramnios or excess amniotic fluid during pregnancy? |
| oligohydramnios | Is oligohydramnios (low amniotic fluid levels) mentioned in the clinical notes? |
| fetal_demise | Do the notes indicate a history of fetal demise or stillbirth in any previous pregnancies? |
| history_of_obstetric_surgery | Is there any mention of prior uterine surgeries such as myomectomy or endometrial ablation? |
| placental_pathology | Do the clinical notes mention any placental pathology or abnormality identified during pregnancy? |
| ethnicity_risk_factor | Do the notes specify the ethnicity of the patient, particularly if they are Asian, African American, or Hispanic? |
| IV_fluid_overload | Is there mention of IV fluid overload or issues with fluid management earlier in the pregnancy? |
| HELLP_syndrome_history | Do the notes mention a history or diagnosis of HELLP syndrome (Hemolysis, Elevated Liver enzymes, Low Platelets) in previous pregnancies? |
| high_MCV | Do the notes mention elevated mean corpuscular volume (MCV > 100 fL), or terms like "macrocytosis"? |
| use_of_uterotonics | Do the notes reference prior use of uterotonics (e.g., oxytocin, misoprostol) during pregnancy management? |
| blood_transfusion_history | Is there any mention of a previous blood transfusion earlier in this pregnancy or in prior pregnancies? |
| abnormal_blood_loss_previous | Do the notes mention any abnormal blood loss in previous pregnancies or deliveries? |
| pre_existing_coagulation_disorder | Do the notes mention any pre-existing coagulation disorders (e.g., hemophilia, von Willebrand disease)? |
| low_hemoglobin_during_pregnancy | Do the notes mention low hemoglobin levels (<10g/dL) during the pregnancy? |
| history_of_uterine_inversion | Do the notes mention a history of uterine inversion in previous deliveries? |
| history_of_postpartum_uterine_atony | Do the notes mention uterine atony in previous deliveries or as a risk factor? |
| use_of_antihypertensive_medications | Do the notes mention the use of antihypertensive medications during pregnancy? |
| abnormal_cervical_dilation_progression | Do the notes mention abnormal progression of cervical dilation during pregnancy or prior pregnancies? |
| fetal_abnormality | Do the notes mention any fetal abnormalities (e.g., congenital heart defects, hydrocephalus) that could complicate delivery? |
| high_maternal_weight_gain | Do the notes indicate excessive maternal weight gain during pregnancy (>40 pounds)? |
| hx_of_incomplete_third_stage_labor | Do the notes mention any history of incomplete placental delivery or retained placenta? |
| hx_of_manual_placenta_removal | Do the notes mention any history of manual removal of the placenta in prior pregnancies? |
| hx_of_operative_vaginal_delivery | Do the notes mention any prior use of forceps or vacuum-assisted vaginal deliveries? |
| hx_of_genital_tract_trauma | Do the notes mention a history of genital tract trauma or injury in previous pregnancies? |
| hx_of_obesity_surgery | Do the notes mention a history of bariatric surgery or other surgeries that could complicate delivery? |
| hx_of_abruption_during_delivery | Do the notes mention any prior incidents of placental abruption during delivery? |
| hx_of_severe_hypertensive_disorders | Do the notes indicate a history of severe hypertensive disorders like eclampsia or HELLP syndrome in prior pregnancies? |
| hx_of_fibroids_or_uterine_surgery | Do the notes indicate any history of fibroids or uterine surgery that may affect delivery? |
| hx_of_neonatal_asphyxia | Do the notes mention any history of neonatal asphyxia or need for neonatal resuscitation in previous pregnancies? |
| hx_of_multiparous_gestation | Do the notes indicate prior multiparous gestations (multiple births) as a potential risk factor for PPH? |
| previous_scar_thickness | Do the notes mention any concerns about uterine scar thickness from previous cesarean sections? |
| hx_of_delayed_cord_clamping | Do the notes mention delayed cord clamping as a part of the delivery process in prior pregnancies? |
| use_of_antiretroviral_medication | Do the notes mention the use of antiretroviral medications (e.g., for HIV) during pregnancy? |
| hx_of_autoimmune_disorders | Do the notes mention a history of autoimmune disorders (e.g., lupus, rheumatoid arthritis) that could complicate pregnancy? |
| abnormal_pelvic_size | Do the notes indicate concerns about abnormal pelvic size or shape that could complicate delivery? |
| hx_of_polycystic_ovarian_syndrome | Do the notes mention a history of polycystic ovarian syndrome (PCOS) as a risk factor? |
| hx_of_hyperemesis_gravidarum | Do the notes mention severe hyperemesis gravidarum or excessive nausea and vomiting in previous pregnancies? |
| maternal_heart_conditions | Do the notes indicate any maternal heart conditions (e.g., arrhythmias, congenital heart disease)? |
| hx_of_uterine_bicornuate | Do the notes mention a history of bicornuate uterus or other congenital uterine anomalies? |
| use_of_narcotics | Do the notes mention the use of narcotics (opioids) during pregnancy for pain management? |
| hx_of_anemia_requiring_transfusion | Do the notes mention any history of anemia severe enough to require blood transfusion in previous pregnancies? |
| hx_of_high_blood_pressure_treatment | Do the notes mention any treatments for high blood pressure (e.g., methyldopa, labetalol) during pregnancy? |
| hx_of_peripartum_hysterectomy | Do the notes indicate any history of peripartum hysterectomy in previous pregnancies? |
| hx_of_difficulty_in_labor | Do the notes mention any history of difficulty in labor or prolonged labor requiring interventions? |
| hx_of_maternal_fever | Do the notes mention any maternal fever during labor or pregnancy? |
| hx_of_spontaneous_preterm_labor | Do the notes mention a history of spontaneous preterm labor in any previous pregnancies? |
